# Beyond Low-Hanging Fruit: A Systematic Approach to Assessing Quality Improvement Needs in Healthcare Practices

**DOI:** 10.1101/2021.07.08.21260218

**Authors:** Sylvia J. Hysong, Varsha Modi, Melia J. Wichmann, Daniel R. Murphy

## Abstract

Quality Improvement (QI) tools abound to help clinicians improve the quality of their care. Such tools, however, assume one knows the process that needs attention. A systematic approach is thus needed for selecting the QI activities that have the greatest impact on quality. This study applies Functional Job Analysis (FJA) to systematically identify areas in preventive care most suitable for high-impact QI efforts. Seven internal medicine practice personnel served as subject matter experts (SMEs) in multiple FJA style focus groups to identify the tasks required to ensure successful execution of eight preventive-care measures. Tasks with the greatest prevalence across measures and highest human error consequence ratings were deemed most suitable for QI. SMEs generated improvement recommendations for the identified tasks via focus groups. Tasks with greatest cross-measure prevalence centered on information gathering; tasks with the greatest potential consequence of error centered on medication management, testing and test follow-up (MMTTF). SMEs reported time, space, staffing, and equipment constraints as fundamental barriers impacting all aspects of primary care. Barriers to successful information gathering included issues with front desk staff and the electronic health record. Barriers to MMTTF included lab workflow and patient-related factors. Improvement recommendations included strategic staffing, technology-based solutions, and added in-house testing capabilities. FJA successfully identified two clear, distinct clinic workflow areas that could benefit from QI initiatives. Although our work is limited by its single-site design, results are consistent with prior multi-site FJA research. Contrary to traditional, disease-specific QI approaches, our work identified a core set of processes that, if optimized, can improve care across multiple clinical conditions, and presented a novel application of a classic HR tool to make more systematic QI decisions.

## Background

Quality Improvement (QI) tools abound to help clinicians improve the quality of their care, such as workflow process maps, Lean/Six Sigma tools, and Statistical Process Control methodologies. Many such tools, however, assume one knows the process that needs attention. Oftentimes, the process to be improved is identified through salience (e.g., it is the pain point of the moment) rather than strategically (e.g., it is likely to have the most impact). A systematic approach is thus needed for selecting the QI activities that have the greatest impact on quality.

## Objectives

This paper presents the application of Functional Job Analysis (FJA)^1^, a task-oriented methodology for scientifically describing work, as a tool to systematically identify areas most suitable for high-impact quality improvement efforts.

## Methods

### Methodology Overview: Functional Job Analysis

FJA uses structured task statements as the basic building blocks of most human resource (HR) interventions (e.g., training, performance management). Generated with the aid of a facilitator by those who do the work, FJA task statements contain the following elements:

- Who performs the task?
- Performs what action?
- Drawing upon what knowledge / source of information?
- Relying on what skills?
- Using what tools, equipment, work aids?
- In order to produce/achieve what? (expected output)

FJA has been used in healthcare in multiple applications including describing the primary care work domain^2^ and assessing QI metric complexity.^3^ We sought to use FJA to systematically identify in an internal medicine group practice the tasks most commonly conducted in the clinic areas (prevalence), and those with the highest consequence of error.

### Participants

Seven members of a large internal medicine faculty group practice (2 physicians, 3 nurses, and 2 front-office staff) served as subject matter experts (SMEs).

### Procedure

Following a modified FJA protocol,^3^ SMEs participated in multiple, facilitated focus groups. They generated statements for the tasks required to ensure successful execution of eight preventive-care measures for which the clinic was accountable: Breast Cancer Screening, Colorectal Cancer Screening, Controlling High Blood Pressure, Hemoglobin A1C Control, Pneumonia Vaccination, Body Mass Index Screening and Follow-up, Tobacco Screening and Cessation, and MyChart Activation (patient enrollment in electronic patient portal). Each task was rated on two complexity dimensions from FJA: human error consequence (the gravity of the consequence ensuing from performing the task incorrectly) and coordinative complexity (complexity of the coordination among staff required to complete the task). Tasks with the greatest prevalence across measures and highest ratings on these dimensions were identified as most suitable for QI. SMEs generated improvement recommendations for the identified activities in a follow-on focus group using affinity diagrams.^4^

## Results

### Task Prevalence, Complexity, and Consequence of Error

SMEs generated a total of 40 tasks required to successfully achieve the target outcomes of 8 quality measures. On average, 67% of the tasks applied to four or more performance measures. Primary care activities with greatest cross-measure prevalence centered on information gathering (e.g., rooming, patient history, reminders); in contrast, activities with the greatest potential consequence of error centered on medication management, testing and test follow-up (MMTTF). There was no task overlap between these two categories of tasks (See Table 1). Futher, information gathering and tasks were significantly more prevalent (M_IG_ = 7.17, SD =.41 M_MMTTF_= 1.5 SD_MMTTF_= .54 *t*_(1)_=20.31, p=.0001) and significantly lower in human error consequence than MMTTF tasks (M_IG_ = 3.5, SD_IG_ =.83 M_MMTTF_= 5.33, SD_MMTTF_= .81 *t*_(1)_=3.84, p=.003). We observed no significant differences in coordinative complexity between IG and MMTTF tasks (M_IG_ = 2.17, SD_IG_ =.75 M_MMTTF_= 2.0, SD_MMTTF_= 0 *t*_(1)_=.54, p=.599).

**Table 1.** Prevalence of tasks appearing in four our more quality measures

| Task | Measure Prevalence | Human Error Consequence | Coordinative Complexity |
| --- | --- | --- | --- |
| *Tasks with Greatest Prevalence Across Quality Measures: Information Gathering* |  |  |  |
| Place formulary medication order. | 2 | 7 | 1 |
| Teach patient how to manage medical condition and prevent complications. | 1 | 5 | 3 |
| Order patient test(s) or treatment(s). | 2 | 5 | 2 |
| Confirm patient need for vaccination and communicate need to specific personnel. | 1 | 5 | 3 |
| Follow-up on the results of a screening test. | 2 | 5 | 2 |
| Identify, document, and encourage patient adherence to proper medication use. | 1 | 5 | 2 |
| *Tasks with Highest Consequence of Error: Medication Management, Testing, and Test Follow-Up* |  |  |  |
| Review patient's current status in chart. | 8 | 2 | 2 |
| Prepare examination room for incoming patient. | 7 | 4 | 2 |
| Obtain preliminary patient information and escort patient to exam room | 7 | 4 | 2 |
| Satisfy "quality sidebar" health reminders currently due for patient. | 7 | 4 | 2 |
| Obtain and record (or update) medical history | 7 | 4 | 2 |
| Update and document health maintenance information for patient | 7 | 3 | 2 |

Note: Prevalence indicates the number of quality measures in which a given task is required to
successfully achieve the measure's target outcome. Human Error Consequence and Coordinative complexity scales each range from 1-8.

### Barriers, Facilitators, and Improvement Suggestions

SMEs reported time, space, staffing, and equipment constraints as fundamental barriers impacting all aspects of primary care. Issues with front desk staff and the electronic health record were identified as barriers to successful information gathering; lab workflow and patient-related factors were identified as barriers to successful medication management, testing and follow-up. Improvement recommendations included strategic staffing (e.g., floating staff), technology-based solutions (e.g., e-scribe) and added in-house testing capabilities (See Figure 1).

**Figure 1.**
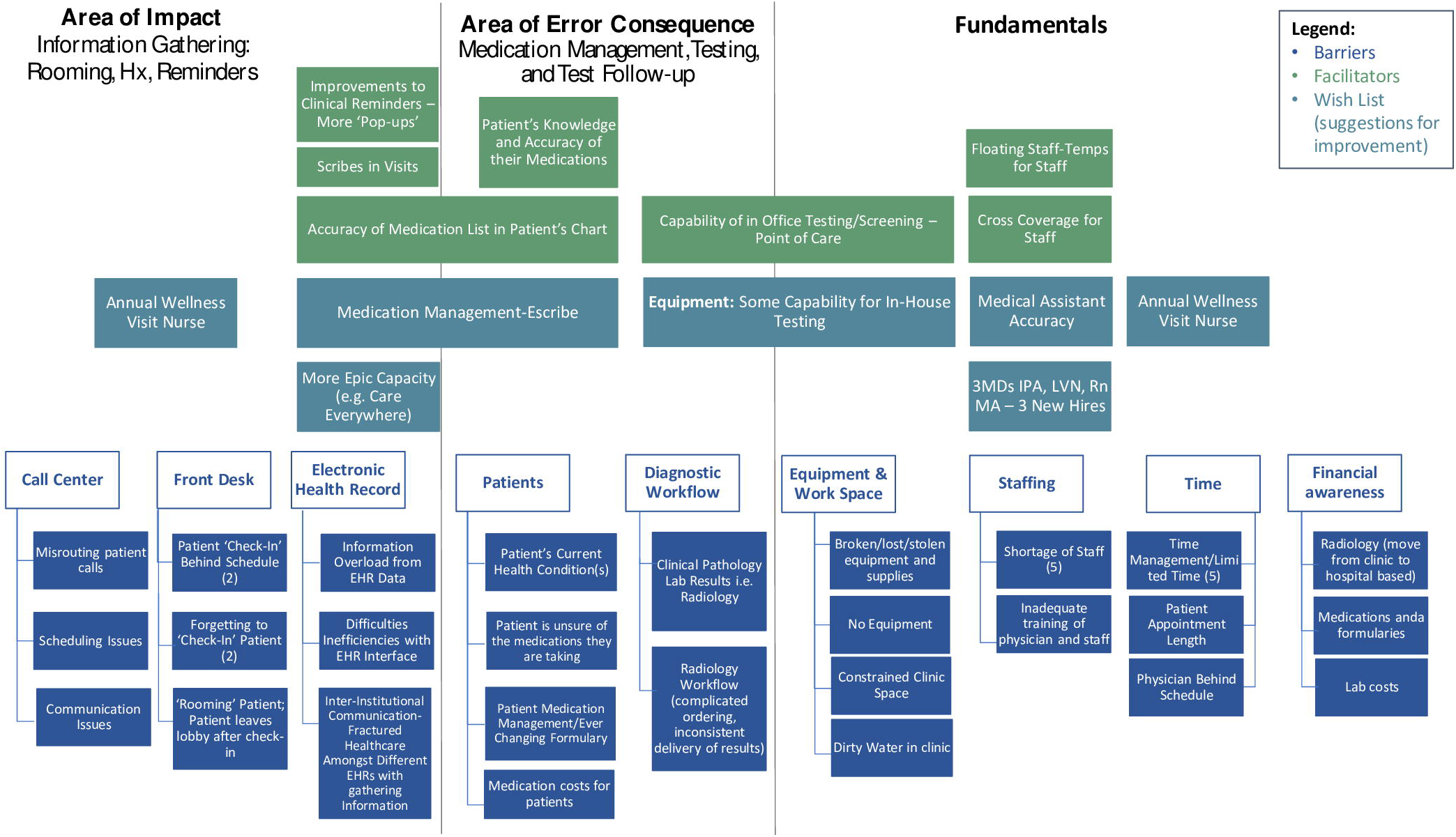
Barriers, Facilitators, and Suggestions for Improving Clinic Workflows in BCM Internal Medicine Faculty Group Practice

## Discussion

As demonstrated by the large number of barriers identified and relatively small number of solutions needed to address them (Figure 1), FJA successfully identified two clear, distinct clinic workflow areas that could benefit from QI initiatives. Contrary to traditional, disease-specific QI approaches, our work identifies a core set of processes that, if optimized, can improve care across multiple clinical conditions. Although our work is limited by its single-site design, results are consistent with prior multi-site research employing FJA to codify primary care work.^2^ Our study thus provides a novel application of a classic HR tool to help make strategic QI decisions.

## Data Availability

Data are available upon written request to the corresponding author at, per institutional policy.

## Notes

### Competing Interest Statement

The authors have declared no competing interest.

### Funding Statement

This work was funded by a Healthcare Innovations Grant from Baylor College of Medicine.

### Author Declarations

Baylor College of Medicine Institutional Review Board protocol # 42846

